# A hemodynamic model to predict regional cerebral blood flow and blood flow reserve in patients with carotid stenosis

**DOI:** 10.1101/2020.07.21.20158840

**Authors:** Joseph P Archie

**Affiliations:** Retired Clinical Professor of Surgery, University of North Carolina, Chapel Hill, Retired Adjunct Professor of Mechanical and Aerospace Engineering, North Carolina State University, Raleigh

## Abstract

**Purpose:** Patients with 50% or greater diameter stenosis are at risk for ischemic stroke due to embolization and/or reduced cerebral blood flow. The hemodynamics of progressive carotid stenosis on cerebral blood flow and blood flow reserve has not been adequately measured or predicted. This information is needed for stroke risk stratification in patients with carotid stenosis. The aim of this hemodynamic model study is to predict the contribution of carotid and collateral blood flows to regional cerebral blood flow and cerebral blood flow reserve in patients with moderate to severe carotid stenosis.

**Methods:** A one-dimensional three-parameter fluid mechanics model for the carotid, collateral and brain vascular systems is used to predict regional cerebral blood flow and blood flow reserve as a function of percent diameter carotid stenosis. The model is based on the principal of conservation of energy as employed by Bernoulli to describe fluid flow on a streamline. When applied to the human cerebrovascular system there are three vascular resistance components; carotid, collateral and brain. Carotid artery vascular resistance is assumed to be a function of fractional percent carotid artery area stenosis. This is not a complex modern computational fluid mechanics study. The model blood flow algebraic equations have simple solutions, one of which gives patient specific collateral resistance values. The solutions are given as patient specific cerebral blood flows and flow reserve as a function of percent diameter stenosis. Established normal clinical values of regional cerebral blood flow, cerebral blood flow auto-regulation and the lower threshold of cerebral perfusion pressure for cerebral auto-regulation are used. Carotid vascular resistance is assumed to be proportional to percent area carotid stenosis. Theoretical solutions use mean systemic arterial pressure of 100mmHg and key clinical values of patient collateral vascular resistance. Clinical solutions use patient measured systemic arterial pressures and carotid stump pressures. The solutions are given as patient specific cerebral blood flow and reserve cerebral blood flow curves over the range of diameter carotid stenosis.

**Results:** Normal regional cerebral blood flow of 50ml/min/100g is predicted to be maintained up to 65% diameter carotid stenosis as reserve blood flow is reduced. With further progression of carotid stenosis to occlusion approximately half of patients are predicted to develop some reduction in cerebral blood flow. However, only about 20% of patients have a decrease in cerebral blood flow below the 30ml/min/100g threshold for cerebral ischemic symptoms.Approximately 10% of patients are predicted to develop regional cerebral blood flow less than the 18ml/min/100g threshold for irreversible ischemic injury. The model predicts critical carotid artery stenosis to be between 65% and 71% diameter depending on mean systemic arterial pressure. With higher degrees of stenosis carotid artery blood flow cannot maintain normal cerebral flow without the contribution of collateral flow. The predicted magnitude of carotid energy dissipation between 60% and 90% stenosis is consistent with observed cervical bruit intensity. Predicted patient specific cerebral blood flow reserve is adequate to prevent significant cerebral ischemia in the majority of patients.

**Conclusions:** Patient specific collateral vascular resistance blood flow curves predict regional cerebral blood flow and blood flow reserve as a function of the degree of diameter carotid artery stenosis. The carotid component of cerebral blood flow is predicted to maintain normal cerebral blood flow up to a critical carotid diameter stenosis of 65% to 71%. Collateral blood flow is necessary to maintain normal cerebral flow at higher degrees of carotid stenosis. The clinical model predicts that many patients do not have sufficient collateral flow to prevent a decrease in cerebral flow should carotid stenosis progress to high grade or occlusion. However, only about 10% of patients are predicted to develop irreversible regional cerebral ischemic injury. Estimated carotid stenosis energy dissipation magnitudes agree with observed cervical bruit intensity. Correlation of predicted cerebral reserve blood flow curves with clinically measured cerebrovascular reactivity/reserve has the potential to predict the probability of future cerebral ischemia in asymptomatic patients with 60% to 80% stenosis.

## Introduction

Extra-cranial carotid artery stenosis and atherosclerotic plaques are a major cause of ischemic stroke. While the majority are caused by emboli from an unstable carotid plaque, reduced regional cerebral blood flow is can be a significant contributing factor or primary cause. Carotid revascularization by endarterectomy is currently the treatment of choice for symptomatic patients with 50% or greater diameter carotid stenosis. Asymptomatic patients with moderate carotid stenosis are generally managed with current best medical therapy and duplex scan surveillance. Carotid reconstruction is often considered when diameter stenosis progresses to 70% or more. In the past decade there have been more carotid endarterectomies performed in the United States on patients with asymptomatic 70% or greater diameter carotid artery stenosis than on patients with symptomatic stenosis. The rational for surgery in asymptomatic patients is the perceived risk of embolization or reduced cerebral blood flow with progression of carotid stenosis or occlusion. Currently there is no accepted method for estimating the risk of cerebral ischemia due to decreased cerebral blood flow if carotid stenosis progresses. Cerebrovascular reactivity measured by pharmacologic cerebral vasodilation has been used to estimate cerebral blood flow reserve. However, there are no methods/techniques to quantitatively predict the amount of regional cerebrovascular flow reserve necessary to protect patients from reduced cerebral blood flow with further progression of carotid stenosis. Early investigations into this problem centered on the concept of critical carotid stenosis. To date no model adequately predicts cerebral blood flow or degree of flow reserve in patients with 70% to 99% carotid stenosis. This includes modern fluid dynamics/hemodynamics computational methods. The aim of this basic fluid mechanics study is to predict the contribution of carotid and collateral blood flows to regional cerebral blood flow and blood flow reserve in patients with carotid stenosis.

## Methods

### Hemodynamic Model, Figures 1 and 2

Figure 1 is a schematic of the three-component hemodynamic model used to predict cerebral blood flow. The components are the parallel carotid vascular resistance, Rc, and collateral vascular resistance, Rw, in series with brain vascular resistance, Rb. While all three vascular resistances are variables, Rw is brain auto-regulated and Rw has patient specific numerical values. Carotid vascular resistance, Rc, is assumed to be a function of percent area carotid stenosis. The mean systemic arterial pressure is Pa, the cerebral perfusion pressure is Pp and the regional cerebral blood flow is Q. This is a one-dimensional three-component basic fluid mechanics model based on Bernoulli’s application of the principal of conservation of energy to fluid flow along a streamline. The concept of vascular resistance energy dissipation was added to the original Bernoulli equations to account for viscous fluid energy loss. The kinetic and gravitational energy losses are negligible relative to the viscous flow energy losses. The input energy, systemic arterial pressure, Pa, is energy per unit volume flow dissipated in the three component resistances. The algebraic equations for Figure 1 are:

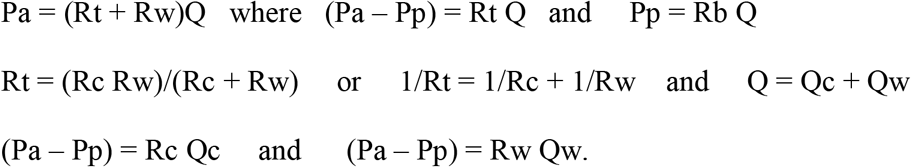

**Figure 1.**
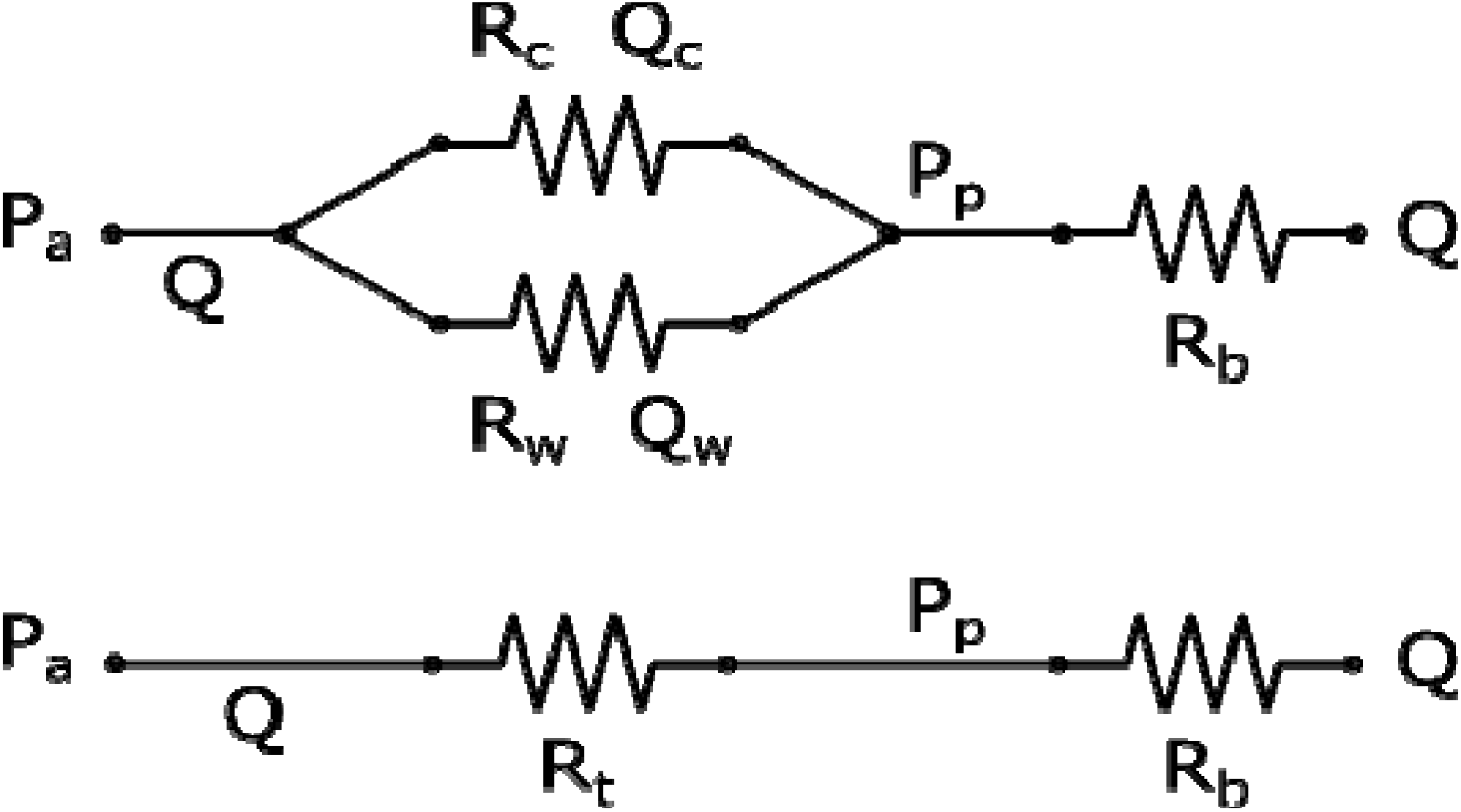
The three-component energy conservation/dissipation model. Pa is mean systemic arterial pressure, Pp is mean cerebral perfusion pressure, Rc is carotid stenosis vascular resistance, Rw is patient specific collateral vascular resistance, Rb is brain vascular resistance, Q is total cerebral blood flow and Qc and Qw are the parallel blood flows that make Q. Rt is the resultant parallel carotid and collateral vascular resistance. As given in the text, Q = Qc + Qw and Rt = RcRw/(Rc + Rw).

**Figure 2.**
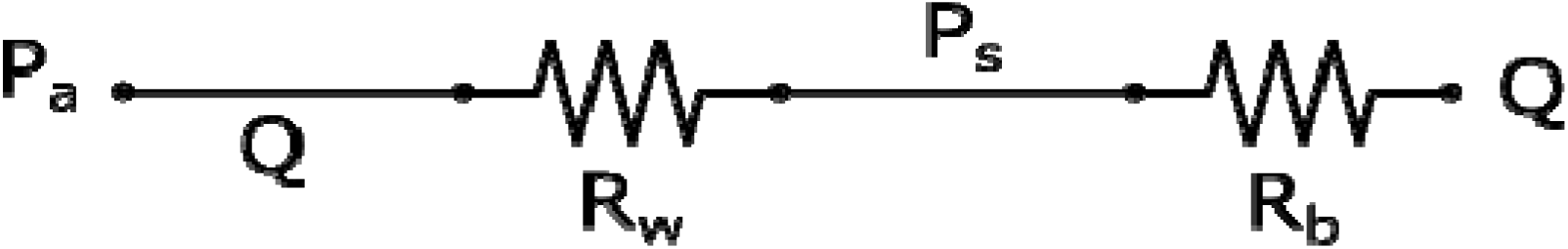
The model given in Figure 1 when there is no carotid blood flow, Rc = ∞, and Rt = Rw. The cerebral perfusion Pp is measued carotid artery stump pressure, Ps, Pt = Ps and the model equation is Rw = Rb (Pa/Ps – 1). When Ps, Pa and Rb are known, the solutions for patient specific Rw values are used in the model to determine cerebral blood flow and cerebral blood flow reserve as a function of percent diameter carotid stenosis for individual patient’s Rw value.

This model is for regional cerebral blood flow in ml/min/per 100 gram brain. This is not a total cerebral hemisphere model, primarily because established human cerebral blood flow values are based on 100grams of brain. Accordingly, carotid and collateral blood flows and all three vascular resistance values are per 100g of brain.

Figure 2 is a special case of the model when the carotid artery is occluded. This occurs clinically with carotid test catheter-balloon occlusion or with temporary carotid clamping during surgery. Carotid stump pressure, Ps, is cerebral perfusion pressure, Pp, when the carotid is occluded and in Figure 2 Pp is replaced by Ps. The carotid resistance, Rc, is absent in Figure 2 because carotid flow, Qc, is zero (Rc = ∞) and Rt becomes Rw. The Figure 2 equations are;

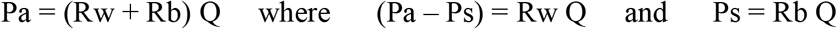

and the solution for Rw, is Rw = Rb (Pa/Ps – 1).

When the values of Pa, Ps and Rb are known the patient specific collateral resistance values, Rw, are determined. With this information the complete model (Figure 1) predicts cerebral blood flow and blood flow reserve for patient specific collateral resistance blood flow curves as a function of percent diameter stenosis.

### Normal Cerebral Blood Flow and Auto regulation Values

Regional cerebral blood flow is normally maintained near 50 ml/minute/100 grams of brain by auto-regulation over mean systemic cerebral perfusion pressures of 50mmHg to 150mmHg (1,2,3). At normal systemic arterial pressures pharmacologically induced maximal cerebral vasodilation has been shown to increase cerebral blood flow two fold in normal humans (4,5). The 50mmHg lower limit of brain auto-regulation cerebral perfusion pressure for normal brain blood flow, Q = 50ml/min/100g means that cerebral vascular resistance Rb, is 1.0 in the model, (Rb = Pp/Q). When cerebral perfusion pressure, Pp, is less than 50mmHg cerebral blood flow is pressure dependent in the model, (Q = Pp/Rb = Pp).

### Carotid Stenosis Resistance, Rc, Figure 3

A key assumption in this model is the non-linear functional relationship between carotid artery stenosis resistance, Rc, and fractional percent diameter carotid stenosis, X; Rc = 1/(1/X^2^ – 1) as given in Figure 3. Carotid artery stenosis resistance, Rc, is defined as the ratio of pressure gradient, (Pa – Pp), to fluid flow, Qc, or Rc = (Pa – Pp)/Qc. This has been studied extensively for carotid artery stenosis. The two classical laminar flow fluid mechanics solutions, Poiseuille’s steady flow and Womersley’s pulsatile flow, account for an arterial pressure gradient of less than 5mmHg at normal carotid artery blood flow rates and even less if flow is decreased (6). When applied to carotid stenosis at normal flow rates both of these two classic laminar flow solutions predict that blood flow becomes turbulent when percent diameter stenosis approaches 50% (75% area carotid stenosis).

**Figure 3.**
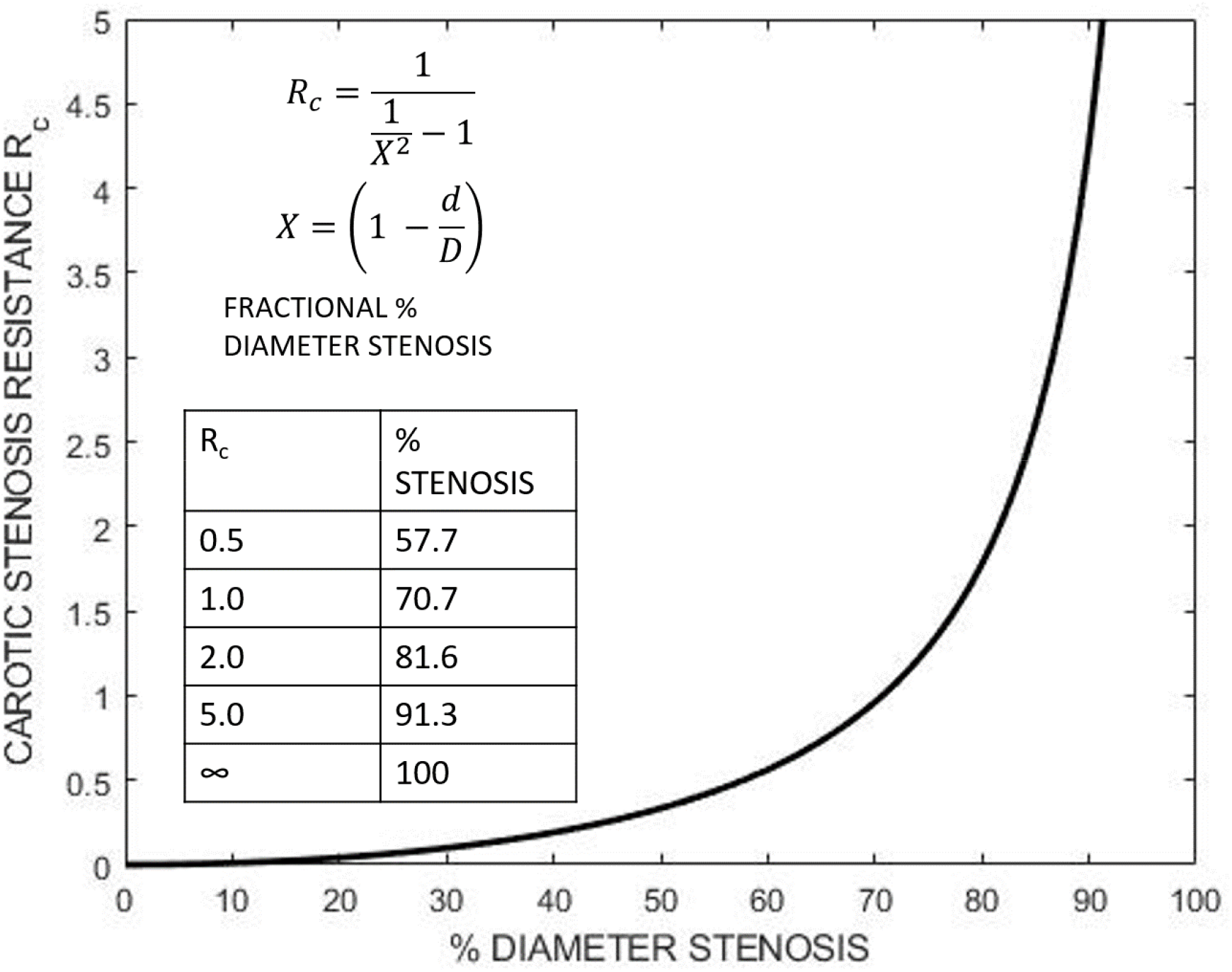
Carotid stenosis vascular resistance, Rc, a measure of viscous blood flow energy dissipation, is assumed to be a function of fractional percent area stenosis, X^2^.

In the equation Rc = 1/(1/X^2^ –l), X is fractional diameter stenosis, defined as (1 – d/D) and fractional % area stenosis is [1 – (d/D)^2^]. The minimum stenosis diameter is d and the normal internal carotid stenosis diameter is D. This function matches the required two endpoints of Rc = 0 when X = 0 (normal carotid) and Rc = ∞ when X = 1.0 (occluded carotid). It correlates closely with the predicted transition to early turbulence at about 50% diameter carotid stenosis and with a decrease in cerebral blood flow above 65% to 71% diameter stenosis in patients with collateral resistance, Rw, greater than 1.0.

### Collateral Vascular Resistance, Rw, and Carotid Stump Pressure, Ps

As presented in Figure 2, patient specific values of collateral resistance, Rw, can be calculated from the model solution Rw = Rb(Pa/Ps – 1). While carotid stump pressure, Ps, is the primary determinant of Rw, both mean systemic arterial pressure, Pa, and cerebral vascular resistance, Rb, contribute. There are a plethora of published carotid stump pressure values ranging from near zero to near systemic arterial pressure. Mean/average values of Ps are reported to be between 40mmHg and 50mmHg and in larger series the median is close to 45mmHg (7, 8). Some studies report systolic rather than mean stump pressures. Systolic stump pressures are over estimates but approach the more accurate mean values at lower pressures due to pressure waveform damping. Stump pressures are used to clinically estimate the adequacy of collateral perfusion to protect the brain during the period of carotid clamping for surgery. The recommended lower carotid stump pressure cut-point values for shunt use range from 18mmHg to 50mmHg. The most commonly used value is 25mmHg, as popularized by Moore (9, 10). The carotid stump pressure values used in the theoretical results of Figures 4a and 4b are, zero, 18, 25, 30, 50 and 75mmHg. In the clinical results given in Figures 5a and 5b the carotid stump pressures are patient measured mean and standard deviation values.

**Figure 4a.**
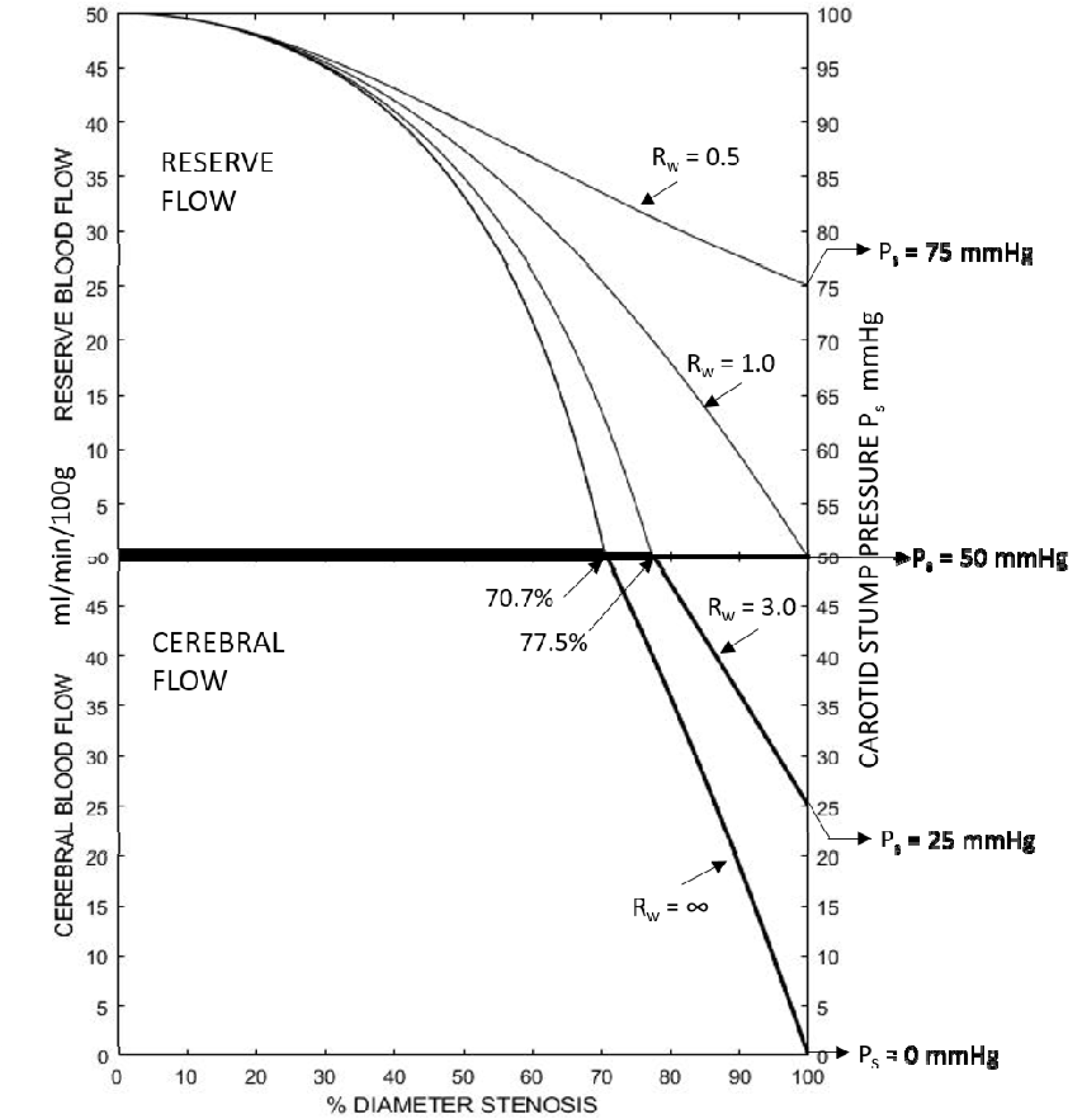
Theoretical solutions for regional cerebral blood flow and blood flow reserve as a function of percent diameter stenosis (Rc = 0 to ∞). The four patient specific blood flow curves are Rw = ∞, Rw = 3.0, Rw = 1.0 and Rw = 0.5. The upper section is flow reserve and lower section regional cerebral blood flow where the normal value is 50ml/min/100g brain. The four Rw curves represent the spectrum of patient values, however Rw = ∞ and Rw = 0 are extremes that do not occur clinically.

**Figure 4b.**
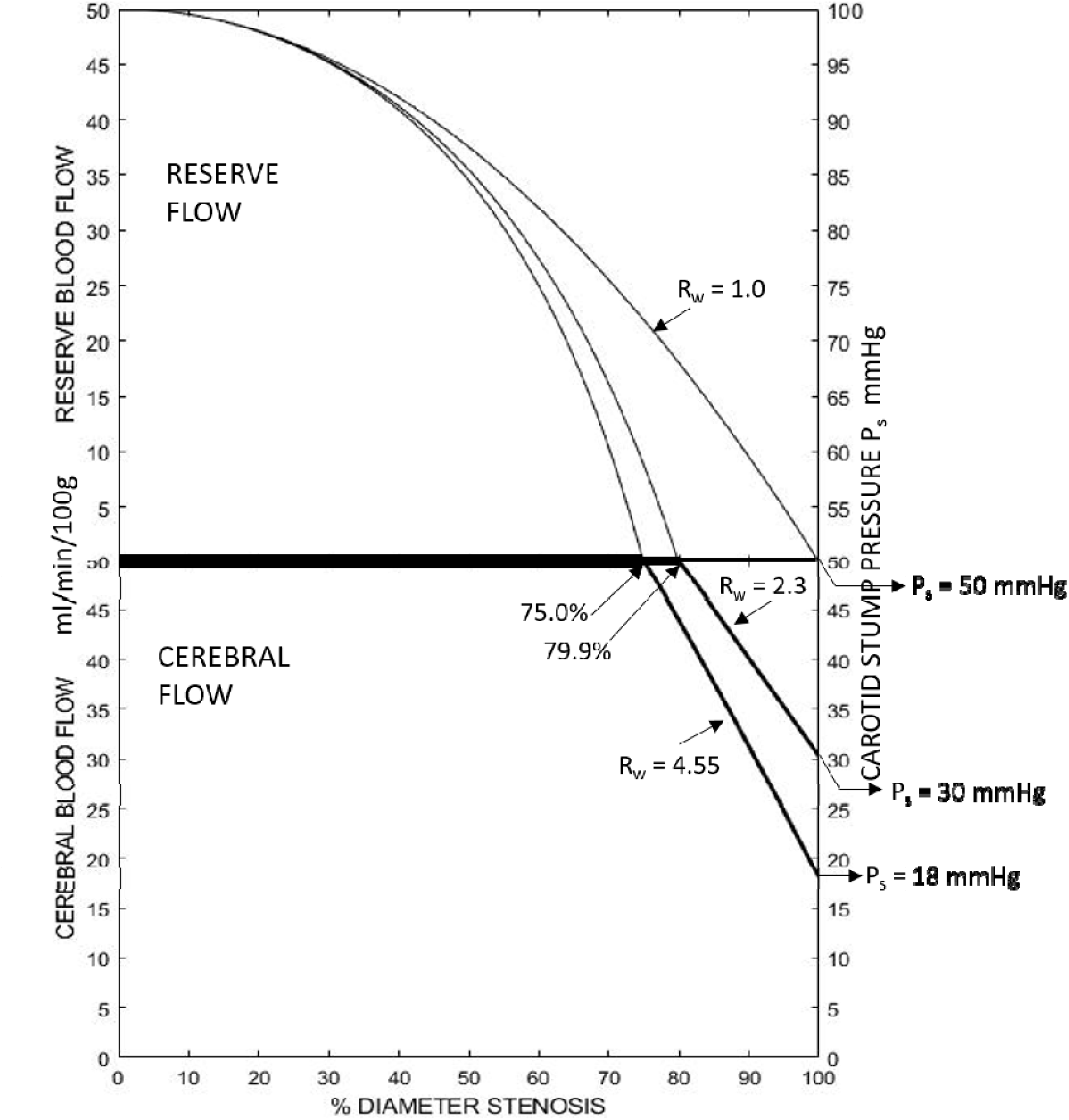
Similar to figure 4a, these are the theoretical blood flow solutions for three clinically important values of collateral vascular resistance. Rw = 4.55 is the solution when carotid stump back pressure Ps, is 18mmHg, the threshold below which cerebral ischemia is irreversible. Rw = 2.3 is when Ps = 25 mmHg, a commonly used threshold below which a shunt is recommended for carotid surgery. Rw = 1.0 is when Ps = 50mmHg, an average patient value.

**Figure 5a.**
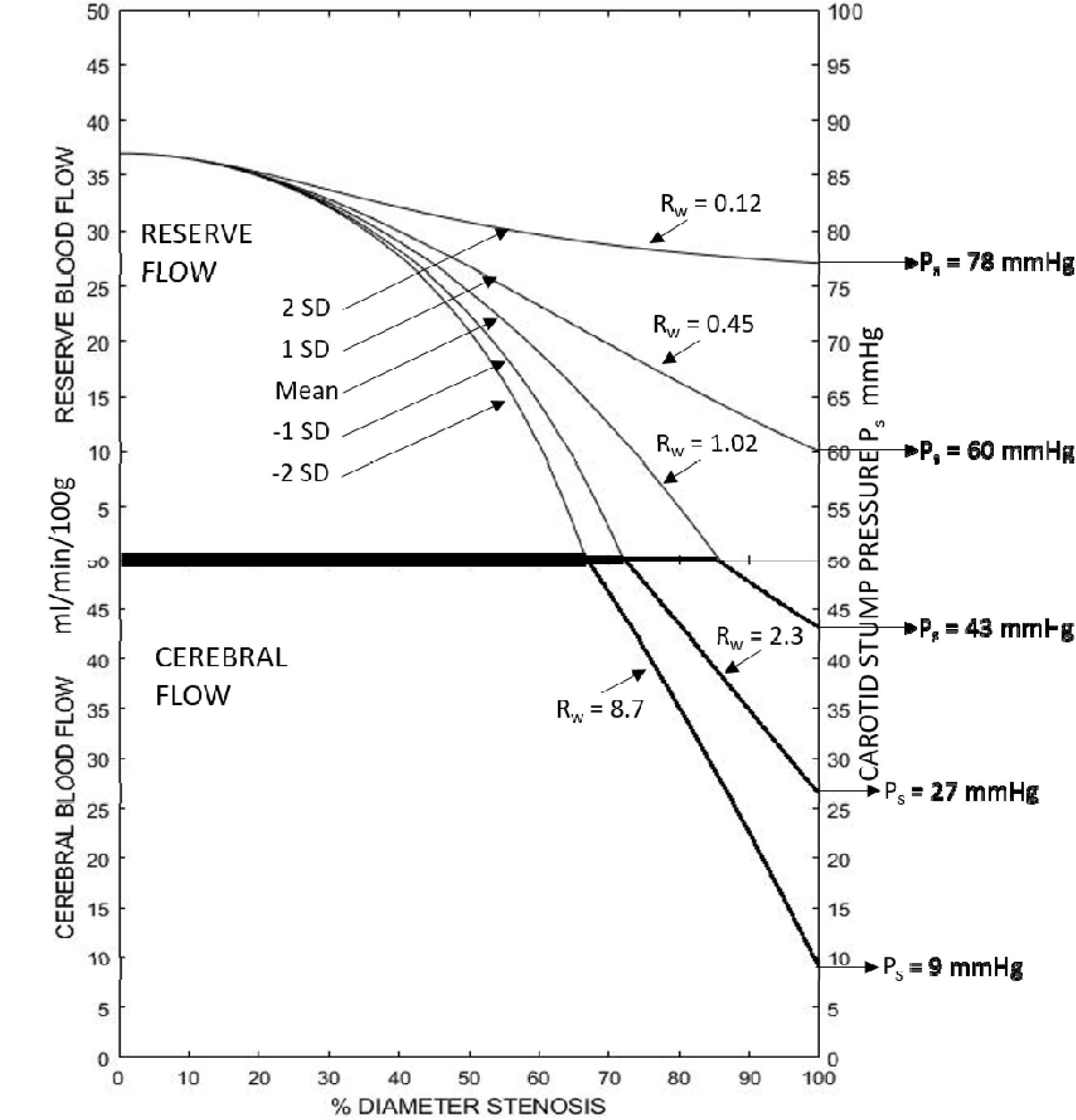
Clinical solutions for regional cerebral blood flow and blood flow reserve as a function of percent diameter carotid artery stenosis. The five patient specific Rw curves are clinically measured carotid stump pressures; mean Ps, mean + ISD, mean + 2SD, mean – 1 SD and mean - 2SD. The mean systemic arterial pressure is 87mmHg.

**Figure 5b.**
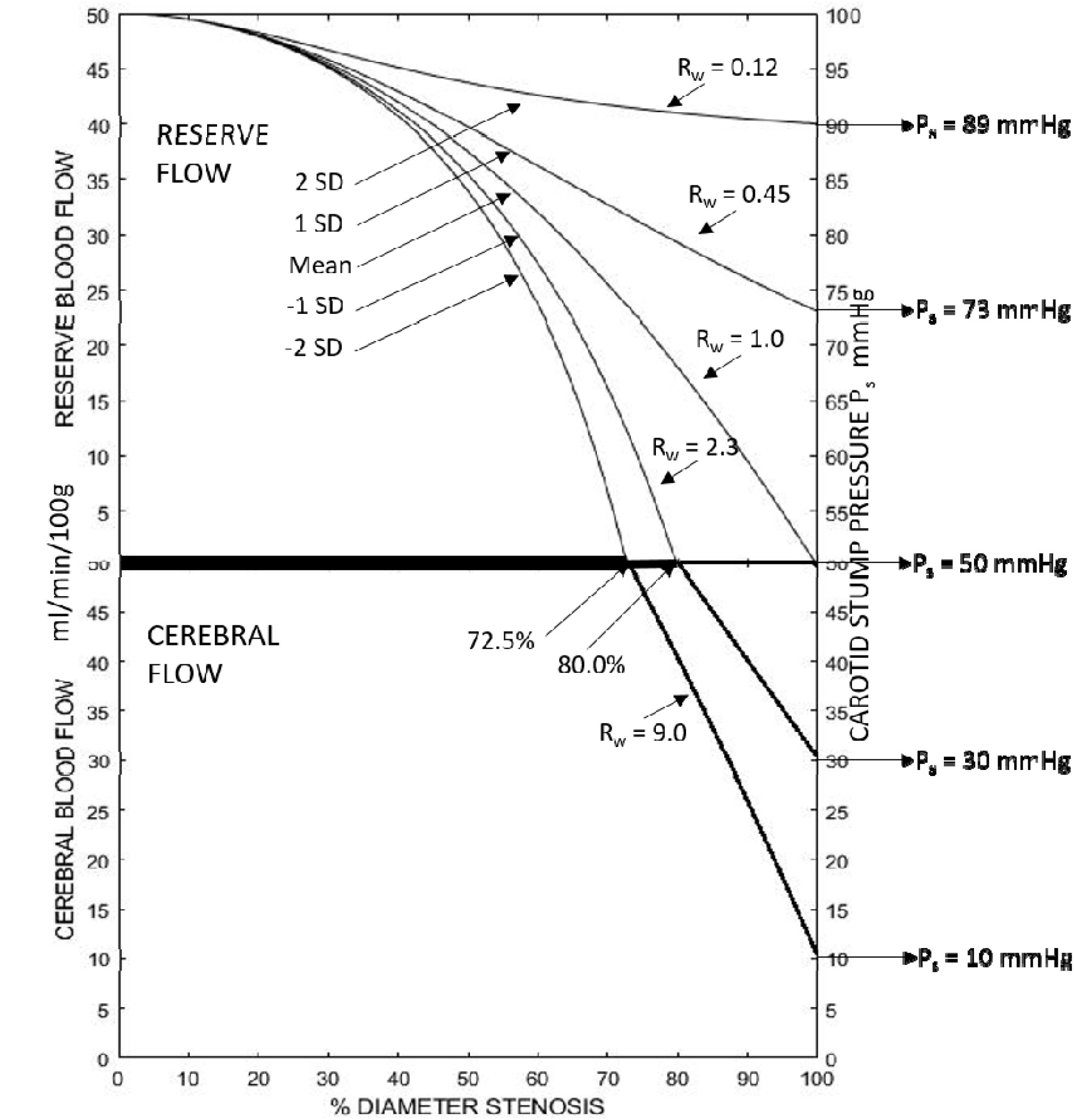
Similar to Figure 5a these are the clinical solutions using mean systemic arterial pressure, Pa, is 100 mmHg The five RW blood flow curves are slightly different because the Figure 2 model solution is Rw = Rb (Pa/Ps - 1). While carotid stump pressure, Ps, is the primary determinant of collateral vascular resistance, systemic arterial pressure, Pa, plays a minor role.

### Cerebral Blood Flow and Reserve Blood Flow Solutions

Solutions to the model equations are obtainable because collateral vascular resistance values, Rw, can be determined and the resulting individual Rw blood flow curves predict patient specific cerebral blood flow and blood flow reserve over the full range of carotid diameter stenosis. Because the lower threshold of cerebral perfusion pressure is 50mmHg, cerebral vascular resistance Rb = 1.0 when Ps < 50mmHg and Rb = 2(Ps/100) when Ps > 50mmHg (Rb = 1.0 to 2.0). As given in Figure 4 and subsequent figures the solutions are non-linear because both brain vascular resistance, Rb, and carotid vascular resistance, Rc are variables.

## Results

### Theoretical Model Solutions, Figures 4a and 4b

The figure 4a solutions are when mean systemic arterial pressure, Pa, is assumed to be 100mmHg, carotid resistance, Rc, varies from zero to infinity (diameter stenosis from 0% to 100%) and collateral vascular resistance, Rw, has 4 specific values. The horizontal axis is 0% to 100% diameter stenosis (0% to 1.0% fractional diameter carotid stenosis, Rc = 0 to ∞). The vertical axis has two sections. The lower section is predicted cerebral blood flow, Q, from 0 to 50 ml/min/100g and the upper section is cerebral blood flow reserve from 0 to 50 ml/min/100g. The four collateral vascular resistance blood flow curves are Rw = ∞, Rw = 3.0, Rw = 1.0 and Rw = 0.5 as given from left to right. The Rw = ∞ curve is cerebral blood flow and blood flow reserve when there is no collateral blood flow, only carotid flow. Both Rw = ∞ and Rw = 0 (not used) are extreme values that do not occur clinically. The Rw = ∞ blood flow curve is the baseline from which the contribution of collateral flow to cerebral blood flow can be compaired. The Rw = 3.0 curve is when Ps = 25 mmHg, a commonly used threshold carotid stump pressure value. Patients with Rw > 1.0 are predicted to develop some degree of reduced cerebral blood flow after carotid stenosis exceeds 70.7%. All carotid and collateral blood flow reserve is utilized at carotid occlusion. The Rw = 1.0 curve is when Ps = 50mmHg, near the average values measured in patients. When Rw = 1.0 all blood flow reserve (both the carotid and collateral systems) is used while maintaining normal blood flow of 50ml/min/100g as Rb auto-regulates from 2.0 to 1.0 and carotid stenosis progresses to occlusion. Patients with collateral resistances less than 1.0, Rw < 1.0, are predicted to maintain normal cerebral blood while decreasing flow reserve with progressive stenosis and maintain some reserve at occlusion.

A useful way to view the solutions in Figure 4a is to consider the far left Rw blood flow curve where Ps = zero and Rw = infinity. All cerebral blood flow and reserve flow is from the carotid artery, that is, no collateral flow. Beginning at zero percent carotid stenosis, cerebral blood flow is normal 50ml/min/100g, Rb = 2.0 and reserve flow is 50ml/min/100g. At 70.7% diameter stenosis cerebral auto regulation/vasodilation is complete, Rb = 1.0 and reserve flow is completely utilized. At 70.7% stenosis cerebral perfusion pressure, Pp, is 50mmHg and further stenosis progression decreases cerebral blood flow, Q, linearly from normal to zero at carotid stenosis occlusion.

In Figure 4a when Rw blood flow curves decrease in value to Rw = 1.0 there is progressively more collateral blood flow but not sufficient to prevent a decrease in normal cerebral blood flow at high degrees of carotid artery stenosis. When Rw < 1.0 normal cerebral blood flow is maintained with high grade stenosis or occlusion and there is both reserve carotid and collateral blood flow. The model predicts that collateral blood flow is essential to prevent cerebral ischemia with high-grade stenosis. Some specifics of this are interesting. With mean systemic pressure, Pa = 100 mmHg and normal regional cerebral flow, Q = 50 ml/min/100g, cerebral blood flow reserve decreases from 50ml/min/100g as the brain auto-regulates from Rb = 2.0 at n zero % stenosis to maximum vasodilation Rb = 1.0. The model equation is Pa = (Rt + Rb)Q = 2.0, and Rt – 1.0. When Rb = 1.0 flow reserve is used up and (Rt + 1.0) = 2.0, or Rt = 1.0. For the Rw = ∞ curve the percent carotid stenosis is predicted to be 70.7%. This may be considered “critical carotid stenosis” when Pa = 100mmHg. From the model equations Rc = (Pa/50 – 1) at critical stenosis. At Rc values (% stenosis values) greater than this cerebral blood flow is predicted to be below normal.

Patients with high collateral resistances (Rw > 1.0) are predicted to be at increased risk of cerebral ischemia with carotid stenosis progression. When patient specific collateral resistance, Rw, is less than 1.0, normal cerebral blood flow of 50ml/min/100g is predicted to be maintained across the full range of carotid stenosis to occlusion and there is some degree of reserve flow depending on the Rw value. To maintain normal flow when Rt is less than 1.0, the brain auto-regulates according to (Rt + Rb) = 2.0. For example if Rt = 0.5, Rb = 1.5 and approximately half of the cerebrovascular reserve is utilized. However, depending on the degree of stenosis and collateral resistance patients fortunate enough to fit into this category are predicted to have adequate cerebral blood flow reserve to prevent cerebral ischemia even if the carotid occludes. In Figure 4b, patient specific curves are given for three clinically important cerebral blood flow values. When Rw = 4.55, Ps = 18mmHg and Q = 18 ml/min/100g, the threshold for irreversible cerebral ischemic injury (11, 12). When Rw = 2.33, Ps = 30mmhg and Q = 30ml/min/100g, the threshold for onset of cerebral ischemic symptoms (13, 14). The blood flow curve Rw = 1.0, an average patient value, also given in figure 4a. The predicted cerebral blood flow and utilization of flow reserve by carotid stenosis when Rw = 1.0 is of interest. When Rc = 1.0, stenosis is 70.7%, Rt = 0.50 and Rb = 1.50. Because Rc/Rw = Qw/Qc (a model solution) Rc= Rw = 1.0, the contribution to regional cerebral blood flow is equal, Qw = Qc = 25ml/min/100g and the model predicts that collateral flow, Qw, does not become predominant until carotid resistance, Rc, exceeds collateral resistance, Rw.

### Model Solutions With Clinical Patient Specific Values, Figures 5a and 5b

These figures are the same format as Figure 4 with regional cerebral blood flow (lower half) and reserve cerebral blood flow (upper half) verses percent diameter carotid stenosis. Clinically measured carotid stump pressures in patients with significant carotid stenosis determine the blood flow curves. The clinical data used are mean systemic pressure Pa = 87 mmHg and mean carotid stump pressure Ps = 43 with a standard deviation (SD) of 17mmHg. These values were calculated from Pa and Ps pressure measurements made during 1,360 carotid endarterectomies over a 15-year period (15). There were 1,133 patients, 621 males, 572 females and 277 bilateral CEA. The ages were 67.5 + 9.0 (mean +/-1SD) years. Indications for carotid endarterectomy were symptomatic (transient ischemia, minor stroke or eye) in 62% and asymptomatic >75% diameter carotid stenosis in 38%. Mean ipsilateral percent diameter stenosis was 80% and mean contralateral stenosis was 31%. This included 98 patients with a contralateral carotid occlusion for which the mean Ps value was 30 mmHg. The occlusion and staged bilateral carotid endarterectomy contributed in part to a wide range of stump pressure values, Ps = 43 +/- 17mmHg. Thus 68% of the 1,360 CEA values fall into the range Pa = 26 to 60 mmHg and 95% between Pa = 9 to 77mmHg. This clinical data set of Pa and Ps values represent the patient population to which this model is aimed. Specifically, all had significant carotid stenosis, were candidates for surgery and were likely to have a similar distribution of co-morbidities. Figure 5a has a measured mean systemic pressure of 87mmHg.

Figure 5b is when systemic arterial pressure is 100mmHg. It illustrates the predicted effect of mean systemic pressure on cerebral blood flow and blood flow reserve. A key feature of this model is being able to calculate collateral vascular resistance, Rw, from clinically measured carotid stump pressure, Ps, and arterial pressure, Pa, measurements. Accordingly, the Rw values used in Figure 5a and b are proportionally spread across a wide spectrum of patients having carotid endarterectomy. Carotid stump pressure, Ps, has been shown to be dependent on systemic arterial pressure, Pa. The ratio Pa/Ps is patient specific (16, 17), a necessity for the model solutions to have clinical value.

### Cerebral Blood Flow Reserve With Moderate to Severe Carotid Stenosis, Figure 6

Figures 5a and 5b illustrate two important model predictions for patients with 60% to 80% diameter carotid stenosis. First, the percentage of patients who will have ischemic symptoms or stroke with carotid stenosis progression to occlusion is low, approximately 15%, as given in the lower portion of the figures. Second, the probability of a patient being in this category is predicted by the amount of reserve cerebral blood flow, as given in the upper part of the Figures.

Figure 6 illustrates reserve cerebral blood flow in patients with 60% to 80% diameter carotid stenosis. The upper section is when patient measured mean systemic arterial pressure, Pa, was 87mmHg and the lower section is when Ps = 100mmHg. The five clinically determined collateral vascular resistance curves, Rw = mean, +/- 1SD and +/- 2SD curves shaded area represents 95% of patients. Those in the lower 1SD zone are predicted to have some level of impaired cerebral blood flow and those in the lower 2SD zone are at high risk of ischemic injury if carotid stenosis progresses to high grade or occlusion. Patients with asymptomatic 60-80% percent stenosis are predicted to have little or no measurable reserve may be considered for carotid revascularization. Patients with mean systemic arterial pressure of 100mmHg in Figure 6 are predicted to have a slightly larger flow reserve than those with Pa = 87mmHg. If this model prediction is clinical valid, preventing mild systemic hypotension in managing blood pressure by this group of patients may be of value.

**Figure 6.**
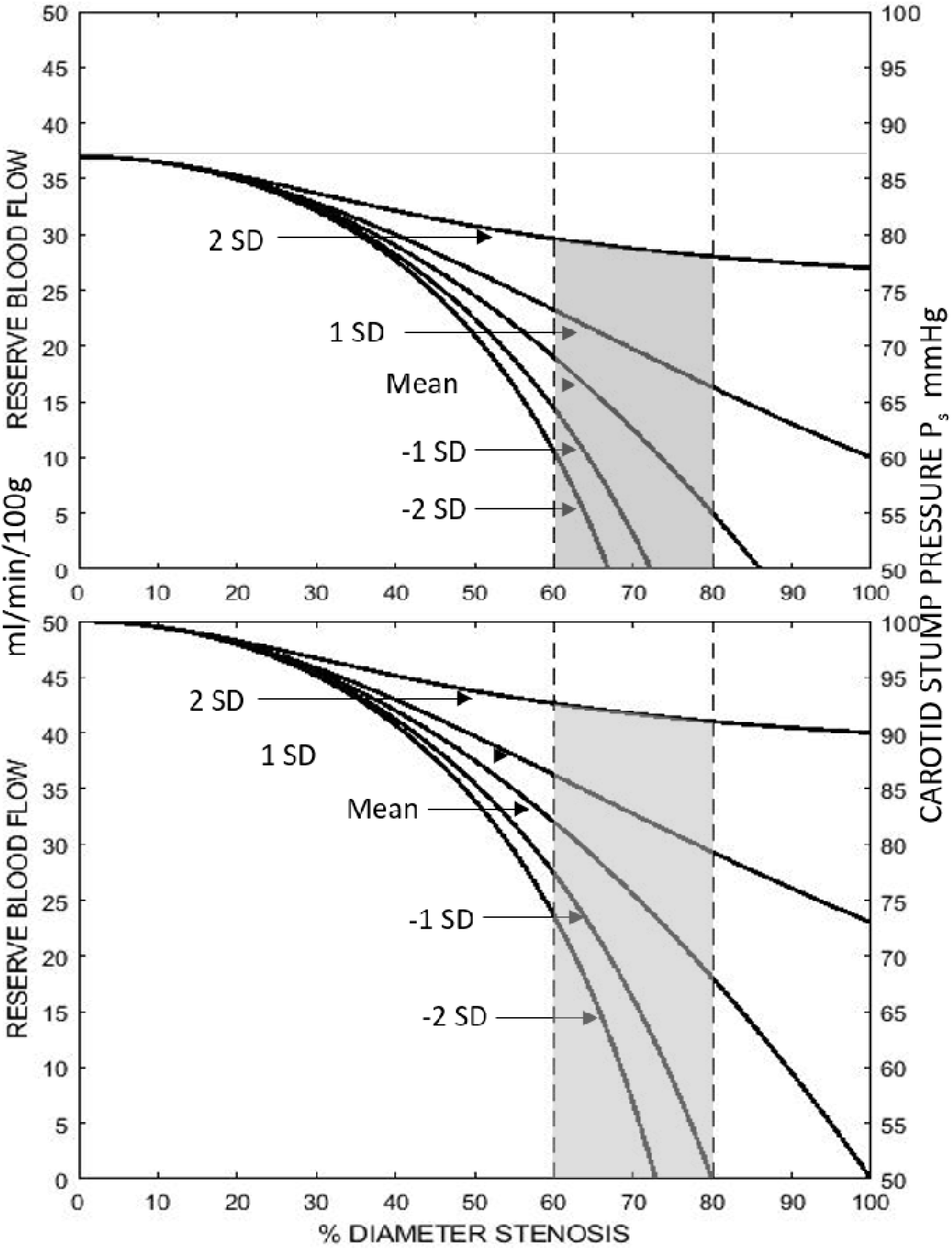
Reserve cerebral blood flow solutions given in Figures 5a and 5b. The shaded areas between 60% and 80% diameter carotid stenosis contains predicted Rw values for 95% of patients having carotid stump pressures measurements.

### Carotid and Collateral Blood Flows, Figures 7a,b,c

These three figures illustrate the parallel contributions of carotid and collateral blood flows to regional cerebral blood flow. The format is similar to that of Figures 4 and 5. Mean systemic arterial pressure, Pa, is assumed to be 100mmHg. On each figure the Rw = ∞ blood flow curve is included as a reference for when all cerebral flow is carotid flow to illustrate the blood flow contribution of the collateral system.

**Figure 7a.**
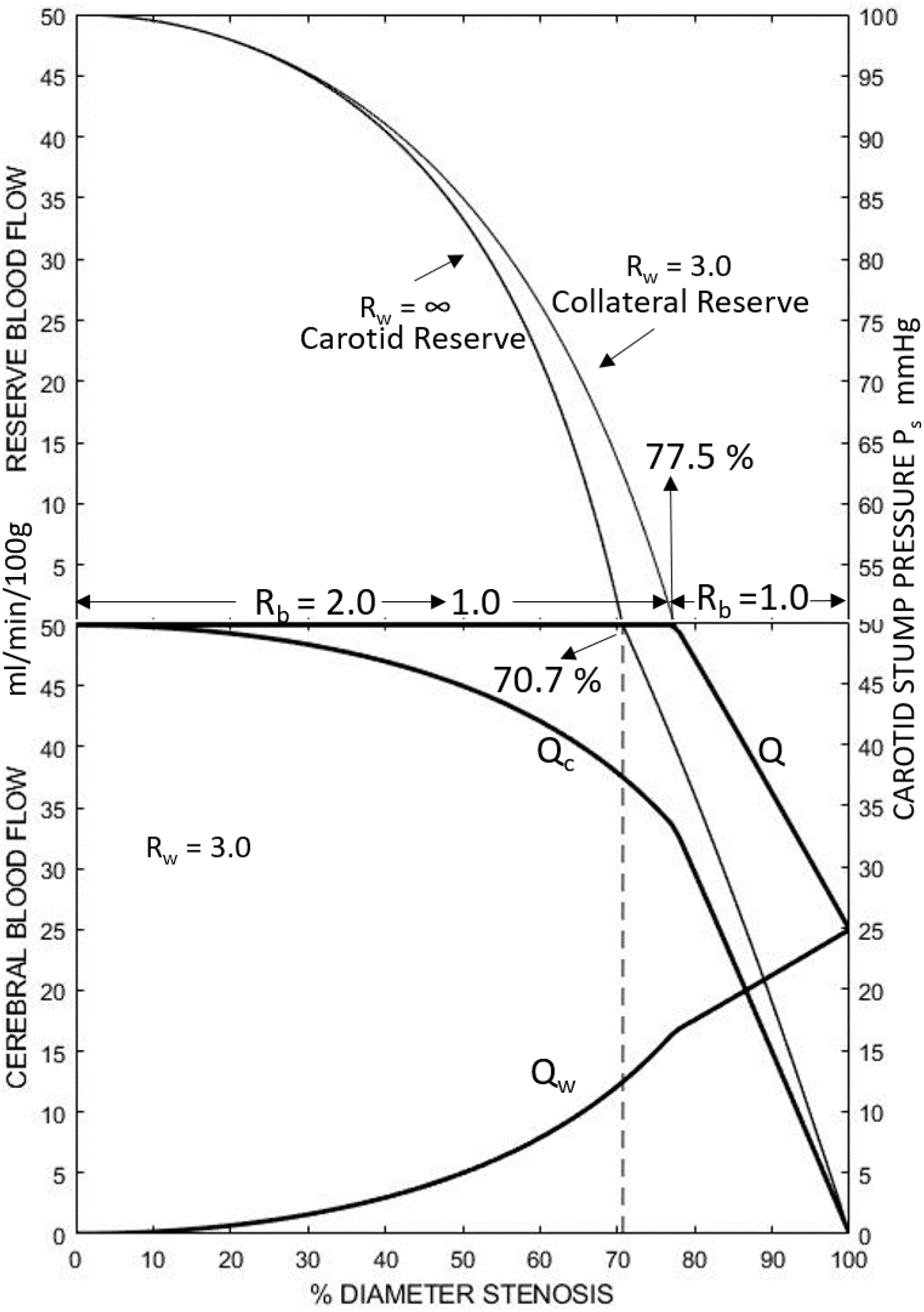
Contribution of parallel carotid and collateral Qc and Qw, to cerebral blood flow versus percent diameter stenosis for the Rw = 3.0 curve. The Rw = ∞ curve is included in all three figures to illustrate the added reserve cerebral blood flow due to the collateral circulation. When Rw>1.0 at some degree of carotid stenosis between 70.7% and 100% cerebral blood flow decreases below normal. In this figure it is 77.8%.

**Figure 7b.**
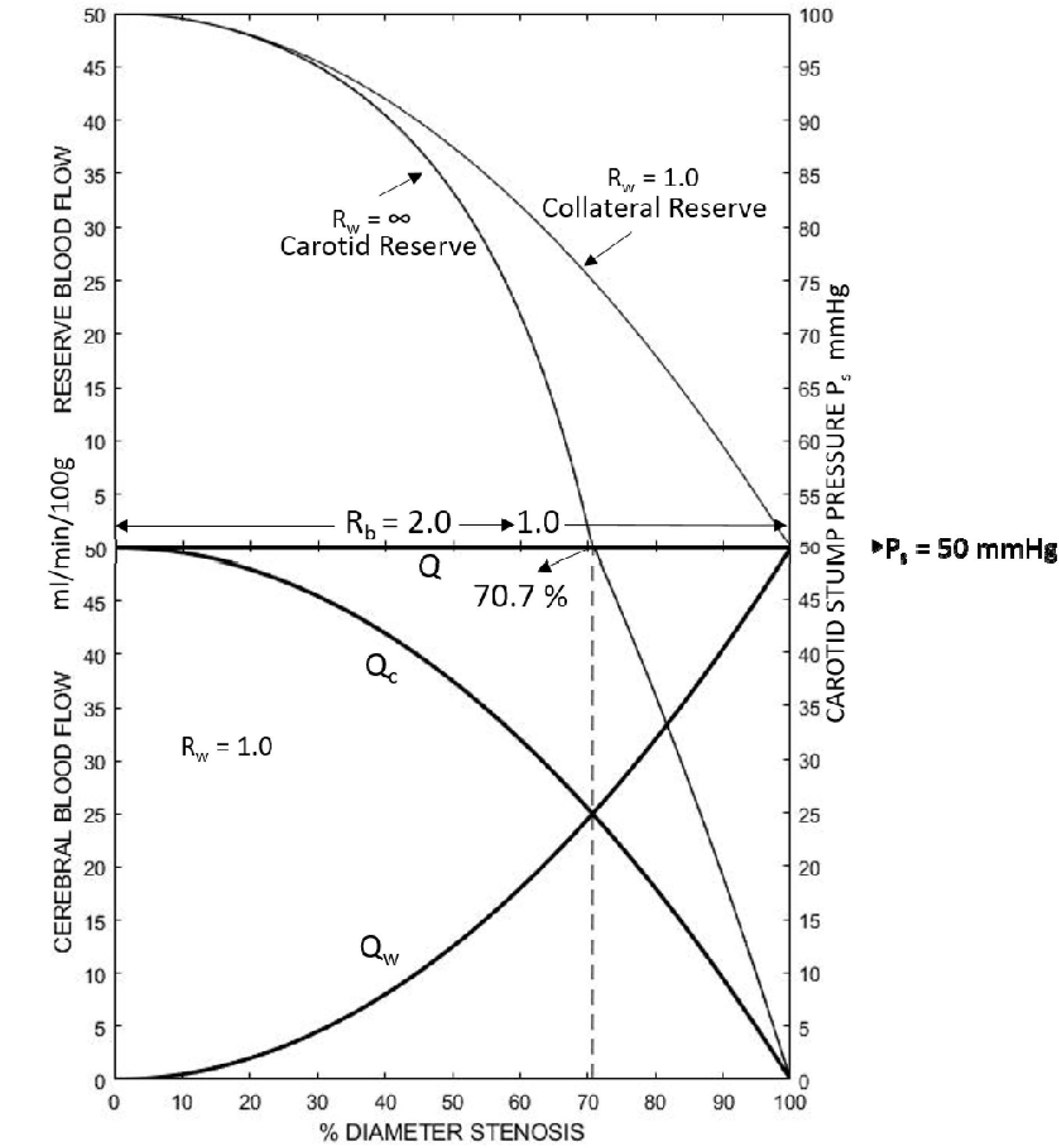
This example of the contribution of the parallel carotid and collateral blood flows, Qc and Qw, to cerebral blood flow is when Rw = 1.0. Normal cerebral flow is maintained at all levels of carotid stenosis as brain auto-regulates from 2.0 to 1.0 and utilizes all carotid and collateral blood flow reserve.

**Figure 7c.**
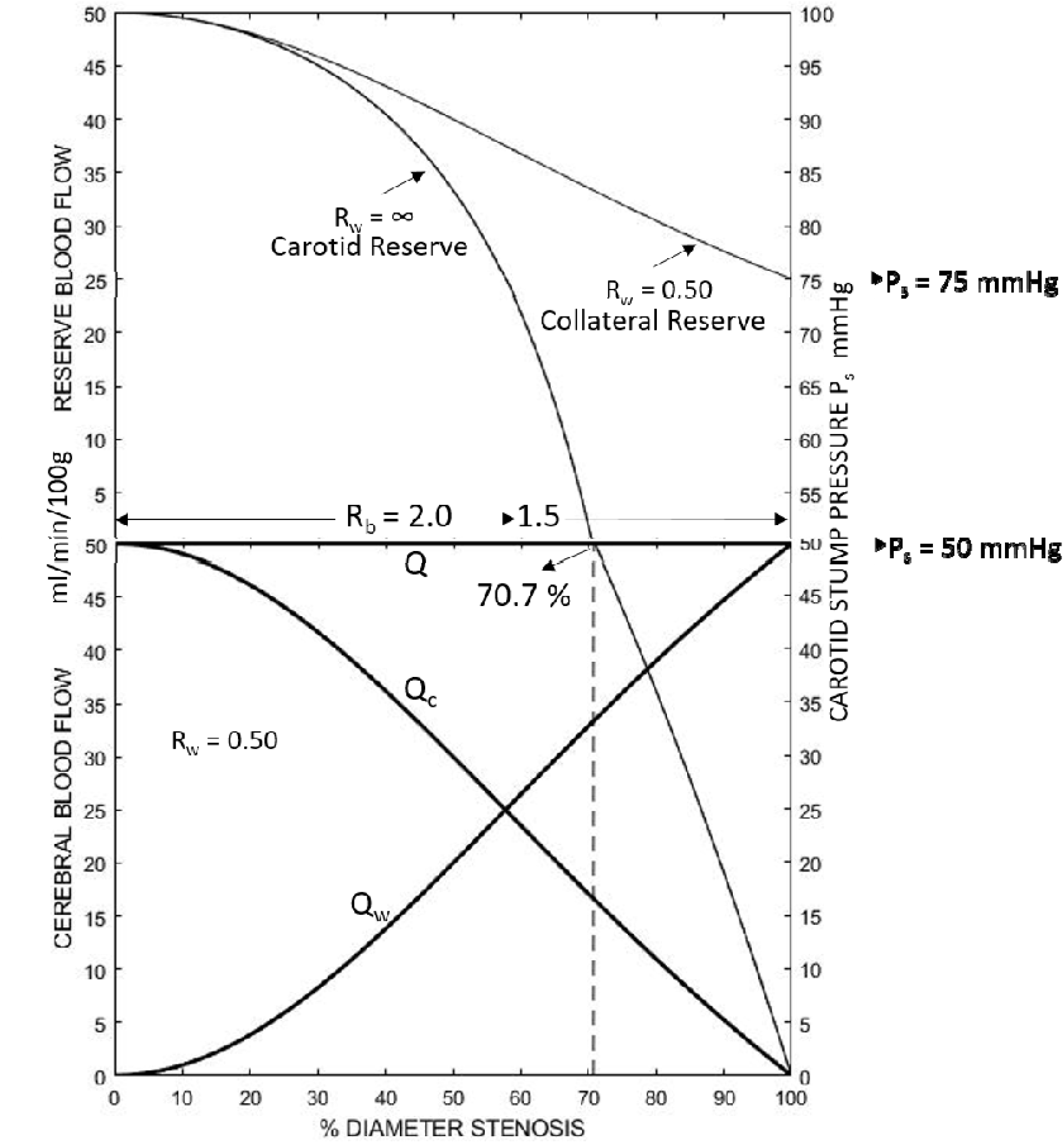
In this example of the contribution of the parallel carotid and collateral blood flows, Qc and Qw, to cerebral blood flow Rw = 0.5. Normal cerebral flow is maintained by brain auto regulation (Rb = 2.0 to 1.5), utilizes all carotid reserve but only part of collateral reserve blood flow.

Figure 7a is collateral resistance Rw = 3.0 and carotid stump pressure Ps = 25mmHg. About 10% of patients undergoing carotid endarterectomy have Ps < 25mmHg and are at risk for cerebral ischemia. Reserve blood flow is utilized completely when carotid stenosis reaches critical 77.5%, (Rb = 1.0). This Rw = 3.0 curve is in the family of patients with specific Rw values greater than 1.0, (Rw > 1.0). In this group reserve flow is completely utilized and cerebral blood flow becomes pressure dependent as carotid stenosis ranges from 70.7% to 100% depending on Rw.

Figure 7b is when Rw = 1.0, the average patient specific value of collateral resistance, consistent with Pa = 100 mmHg and Ps = 50mmHg. The added collateral flow is sufficient to maintain normal cerebral blood flow over the full range of carotid stenosis. Reserve flow is completely utilized at carotid occlusion, Rc = ∞ and Rb = 1.0.

Figure 7c is when Rw = 0.5, Ps = 75mmHg and Rb ranges from 2.0 to 1.5. Patients with collateral resistance values less than 1.0 (Rw < 1.0) are predicted to have adequate cerebral blood flow to prevent ischemia and reserve. In this figure Rw = 0.50 represents a patient at the upper favorable end of the collateral flow spectrum. In all three of these figures there is collateral flow (Rw < ∞) and the parallel carotid and collateral flows mix to give total blood flow curves Rw = 3.0, Rw = 1.0 and Rw = 0.5 respectively.

### Critical Carotid Stenosis, Figure 8

The term “critical carotid stenosis” can mean the degree of stenosis at which cerebral blood flow decreases below normal or when a decrease in cerebral blood flow produces transient ischemic symptoms or stroke. The degree of carotid stenosis is usually given as percent diameter or percent area. These two definitions are based on the anatomically assumption that carotid artery stenosis is a symmetrical reduction in diameter or area of the circular cross section at maximum stenosis, like a “donut”. Many published studies addressing the concept of critical arterial stenosis are biased because they do not account for collateral flow (18). This author was similarly guilty of failing to consider collateral flow in a clinical study of carotid stenosis (19). However, the results from this older study are of value for validating the model used herein. Internal carotid blood flow was measured intra-operatively before and after carotid endarterectomy. Carotid blood flow began to decrease from normal at about 60% - 65% diameter stenosis, as illustrated by the regression lines in Figure 8 (Figure 1 in reference 19). This percent diameter critical stenosis cut-point may be slightly low because some patients in this study had mild early post carotid endarterectomy cerebral hyper-perfusion. Many patients in this older study, as well as in those in other critical carotid stenosis studies probably had adequate collateral blood flow to maintain normal cerebral blood flow at high levels of carotid stenosis or occlusion because of adequate collateral flow. The effect of the absence of collateral flow is illustrated in Figures 4, and 7 by the Rw = ∞ curve, the model blood flow prediction when there is no collateral blood flow contribution. The Rw = ∞ curve intersects normal cerebral blood flow at 65.2% when clinically measured mean systemic pressure was 87mmHg and 70.7% diameter stenosis when mean systemic arterial pressure was assumed 100mmHG. These values are close to the 60% - 65% clinical critical stenosis value in Figure 8. The theoretical model results in Figure 4 and the clinically calculated results in Figure 5 predict that almost half of patients undergoing carotid endarterectomy will not develop a decrease in cerebral blood flow with carotid stenosis progression to occlusion. Further, less than 20% are predicted to have cerebral blood flow less than the clinical symptoms cut-point of 30ml/min/100g (13, 14) and about 10% of patients are predicted to have cerebral blood flow below the clinical stroke cut-point of 18 ml/min/100g (11, 12). The percent diameter stenosis at which cerebral blood flow is predicted to decrease from normal ranges from 65% to 100%. This occurs as Rw increases from zero to 1.0 in Figure 4 and 1.03 in Figure 5. These model solutions an answer to the question; “what is a critical carotid stenosis?”. It is determined by the availability of ipsilateral collateral blood flow as determined by collateral vascular resistance, Rw. In the absence of collateral flow it is predicted to be 65% to 71%.

**Figure 8.**
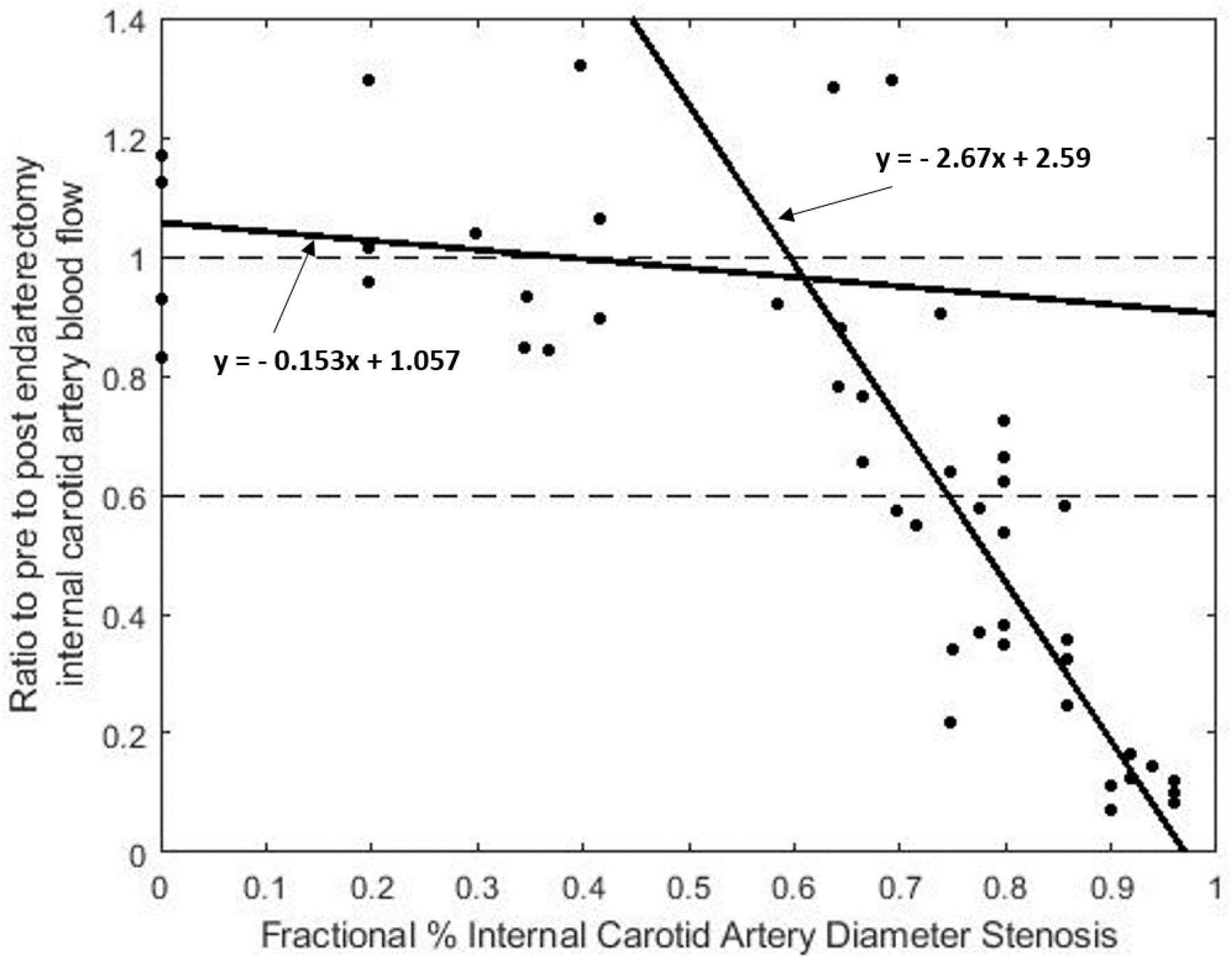
Patient measured carotid blood flow before and after carotid endarterectomy versus percent diameter carotid stenosis from reference #19 Figure 1. As predicted by the model, beyond 60% to 65% diameter carotid stenosis the carotid component of blood flow, Qc, is insufficient to maintain normal cerebral flow.

### Model Energy Analysis and Carotid Bruits

This cerebral hemodynamic model is based on a modification of Bernoulli’s application of energy conservation to fluid flow systems along a streamline. To model the complex human cerebrovascular system that includes carotid stenosis, it is necessary to account for turbulence when diameter carotid stenosis exceeds approximately 50%. The model predictions presented herein are based on the relationship between carotid stenosis vascular resistance and fractional percent diameter stenosis given in Figure 3. The input energy, E, by definition is the product of mean systemic arterial pressure, Pa, and total blood flow, Q, or E = PaQ, and E = (Rt + Rb)Q^2^. This is actually energy per unit time or power. The energy dissipation has two components; RtQ^2^ and RbQ^2^. Carotid stenosis energy is produced by turbulent flow and is dissipated as heat and tissue deformation localized to the region of stenosis in the neck. In contrast, both collateral and cerebral blood flows normally maintain laminar blood flow because the viscous energy is dissipated throughout their vascular beds (6). The energy distribution between the three model components changes significantly as carotid stenosis progresses from zero (Rc = 0) to occlusion (Rc = ∞). This is illustrated using mean arterial pressure Pa = 100 mmHg and normal cerebral blood flow Q = 50 ml/min/100g, giving an input energy PaQ = 5,000 mmHg/ml/min/100g. An easy way to understand the progressive energy dissipation in the three model systems with increasing degree of carotid stenosis is to consider no collateral blood flow, Rw = ∞. At zero carotid stenosis, Rc = 0, the cerebral perfusion is systemic pressure and Rb = 2.0. The model equation is Pa = RbQ when Rb = 2.0 and the total input energy E = 2(50)^2^ = 5,000 units. With carotid stenosis progression to 70.7% stenosis (Rc = 1.0) brain energy utilization decreases to E = 2,500. The 5,000 energy balance is maintained by carotid stenosis energy utilization increasing from zero to 2,500. The 2,500mmHg/ml/min/100g brain carotid stenosis energy loss at 70.7% stenosis is the maximum predicted for carotid stenosis. The progressive increases in carotid energy loss, E, as stenosis increases are Rc = 0.1 (32.2%) and E = 250, for Rc = 0.2 (40.8%) and E = 500, for Rc = 0.35 (50.1%) and E = 875 and for Rc = 0.50 (57.7%) and E = 1,250. As given in Figure 4, Rw = ∞, carotid energy dissipates as cerebral blood flow decreases. When Rc = 3.0, Q = 25 and the total energy is PaQ = 2,500. This is RcQ^2^ = 1,875 carotid and RbQ^2^ = (1) (625) = 625 brain energy loss. With inclusion of collateral flow the maximum carotid stenosis energy dissipation per 100 gram brain decreases from 2,500 units at 70.7 % stenosis in proportion to increasing collateral flow. This is illustrated in Figure 7a for Rw = 3.0. The more collateral flow the sooner carotid blood flow decreases and carotid energy dissipates.

The energy threshold for development of turbulent flow is estimated to be about 250 - 300 based on Rc = 0.33 at 50% stenosis. The energy threshold for the 5mmHg pressure gradient predicted for transition to turbulence by classical laminar flow solutions (6) is about 250 units Q(Pa – Pp) = 50(5). The predicted carotid stenosis energy loss of 2,500 at 57.7% stenosis exceeds this, indicating that the model predicts development of carotid turbulence between 50% and 60% stenosis. However, the large variance in energy dissipation predictions when collateral flow is included may explain to some extent the lack of correlation of cervical bruit intensity with greater than 60% diameter stenosis. Systolic carotid bruits are reported to be present at 50% to 60% stenosis, become continuous with more intensity at greater than 60% stenosis and diminish in intensity as stenosis approaches 90% (20).

The prediction that collateral flow primarily maintains normal cerebral blood flow at high grade carotid stenosis may partially explain the difficulty of obtaining carotid Doppler velocity values that correlate well with high degrees of stenosis. Perhaps an optimal carotid duplex scan is one that combines accurate stenosis imaging with velocity to predict the carotid flow contribution to cerebral blood flow.

## Discussion

The value of hemodynamic models is to give guidance and insight into complex cardiovascular problems. The output solutions are determined by the model design, the input information and the assumptions. The model equations keep track of the variables. The key input data are patient specific values of mean systemic arterial pressure (Pa) and mean carotid artery stump pressure (Ps) from which the model generates patient specific collateral vascular resistance values (Rw). The second key model input is the assumed functional relationship between carotid vascular resistance (Rc) and percent diameter carotid artery stenosis. The additional input data are three; clinically established normal regional cerebral blood flow of 50ml/min/100g, the lower cerebral perfusion pressure threshold for blood flow auto-regulation of 50mmHg and the two fold cerebral blood flow auto-regulation range at normal systemic blood pressure. The model’s output solutions are regional cerebral blood flow and blood flow reserve as a function of percent diameter stenosis, as given by patient specific collateral vascular resistance blood flow curves (Rw). The solutions may have value for predicting cerebral ischemia due to reduced blood flow in asymptomatic patients with progressive moderate to severe carotid artery stenosis. Non-invasive measurements of a patient’s cerebral blood flow reserve or cerebral reactivity and their percent diameter carotid stenosis give a set of coordinates (y = % reserve flow, × = % stenosis) on the model reserve blood flow solution curves that will predicted collateral vascular resistance, Rw. Further, the model predicts critical carotid stenosis at 65% to 71%, that percent area carotid stenosis is a valid assumption to use for carotid artery vascular resistance and that the intensity of predicted carotid blood flow energy dissipation, measurable with carotid bruit acoustics, may inversely correlate with the degree of collateral blood flow. These topics are discussed in the above order.

### Carotid Stump Pressure and Collateral Vascular Resistance, Rw

Carotid stump-back pressure, Ps, is cerebral perfusion pressure, Pp, when the carotid artery is occluded. Carotid stump pressure is considered an estimate of collateral cerebral blood flow with carotid artery occlusion. The hemodynamic model solution for collateral vascular resistance is Rw = Rb (Pa/Ps - 1), where Rb is cerebral vascular resistance, Pa is mean systemic arterial pressure and Ps is carotid stump pressure. For collateral vascular resistance to be patient specific the ratio Pa/Ps must also be patient specific. For example if Pa = 90mmHg and Ps = 45 mmHg, Pa/Ps = 2.0, if Pa is increased to 120 mmHg, Ps should increase to 60mmHg. Clinical evidence of the invariance of the patient specific Pa/Ps ratio was obtained by sequential measurements of both Pa and Ps while increasing systemic arterial pressure as given in Figure 9 (Figure 3 in reference 21). Two other studies report Pa/Ps of 2.05 and 1.90 as mean values in patients with significant carotid stenosis (15, 20). These clinical results support the requirement collateral vascular resistance, Rw, is patient specific for the collateral blood flow component of model solutions in Figures 4 and 5. The third variable necessary to obtain patient specific Rw values is cerebrovascular resistance, Rb. When cerebral perfusion pressure is less than 50mmHg Rb = 1.0 and the solution is Rw = (Pa/Ps -1). When Pp is greater than 50mmHg the solution is Rw = (Ps/50)(Pa/Ps -1).

**Figure 9.**
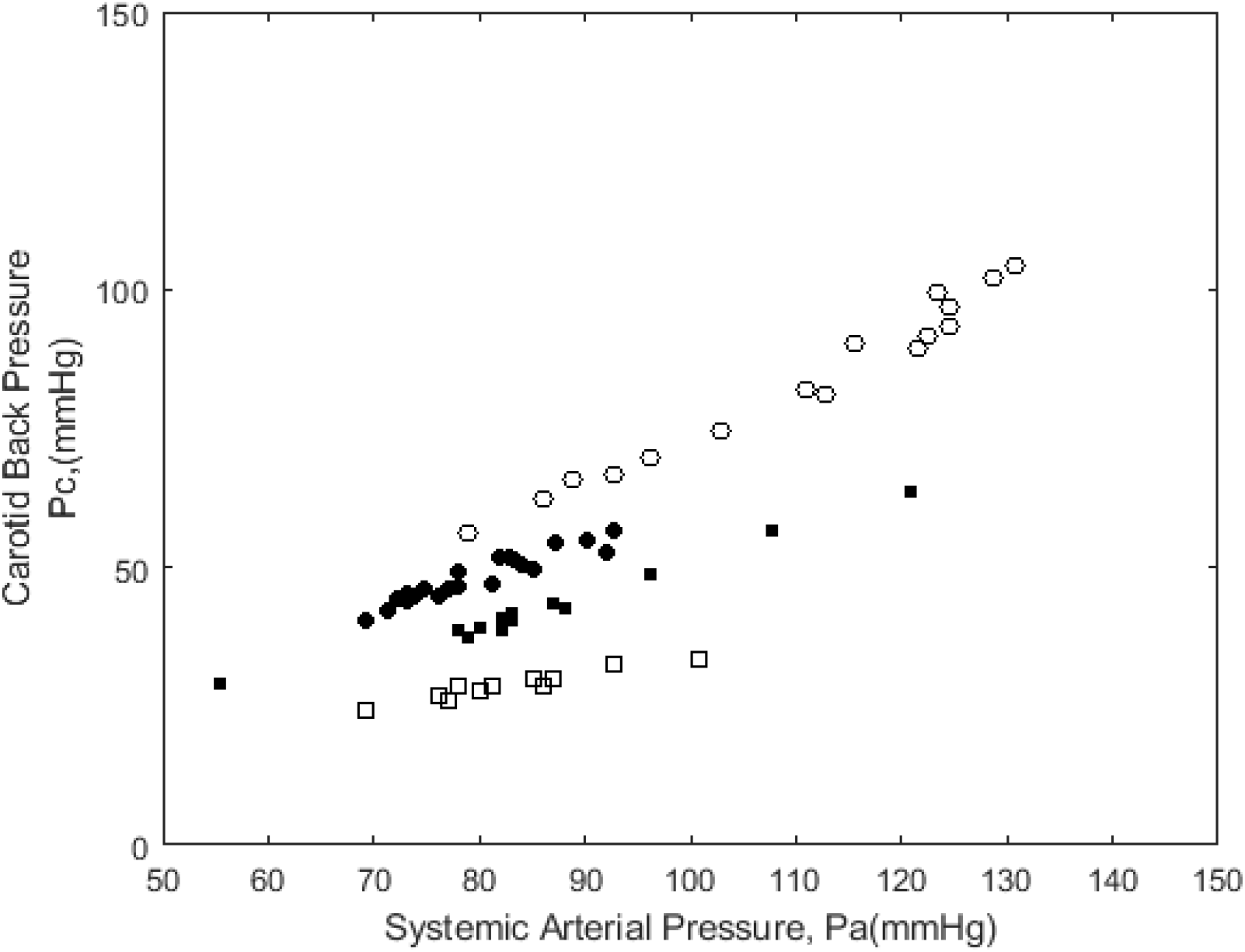
A key component of the model is the necessity for the ratio of mean systemic arterial pressure to mean carotid stump pressure, Pa/Ps, to be within patient invariant, that is, to be between patient specific. The invariance of the slope, Pa/Ps, in these four patient over a range of Pa values illustrate this (Figure 3 in reference #21).

Mean systemic arterial pressure has a small but real effect on the predicted Rw blood flow curves. Comparing the model results in Figure 4, Pa =100mmHg to those in Figure 5a, Pa = 87mmHg illustrates this. The patient specific curve for Rw = 1.0 predicts normal cerebral blood flow over all ranges of carotid stenosis in Figure 4 but in Figure 5a flow is decreased up to 7ml/min/100g with carotid occlusion. The model requires that both Pa and Ps be known to predict cerebral hemodynamics.

### Carotid Stenosis and Carotid Vascular Resistance, Rc

The assumed functional relationship between carotid vascular resistance, Rc, and fractional percent carotid stenosis, X, as given in Figure 3 is a key model input. The assumption that arterial stenosis resistance is proportional to percent area stenosis is an old concept, but to my knowledge it has never been confirmed for carotid stenosis. It implies that the energy dissipation in a circumferential carotid artery stenosis per unit volume flow is proportional to area carotid stenosis. The equation is Rc = 1/ [(1/X^2^) -1], where X = (1 – d/D) is the fractional percent diameter stenosis, where d the stenosis diameter and D the normal carotid artery diameter. It satisfies the requirement that Rc = 0 when percent diameter stenosis X is zero and when Rc = ∞ (carotid occlusion) X = 1.0. This simple assumption generated model solutions consistent with clinical findings, without having to introduce a fudge factor such as a “relativity constant” to make it fit. The solutions are consistent with clinical observed onset of turbulent flow, critical carotid stenosis cut-points and cervical bruit intensity. At 50% to 60% diameter stenosis the model predicts carotid stenosis energy losses exceeding 250 – 300 units consistent with development of turbulent blood flow and the onset of a cervical bruit. Bruit intensity correlates positively with energy losses, which can reach 2500 units. The cardiac cycle distribution of bruit intensity is not predicted because pulsatile flow was not used.

The model predicts critical carotid stenosis at 70.7% (Pa = 100mmHg) or 65.2% (Pa = 87mmHg) consistent with the 60% – 70% value measured clinically in Figure 8. Above 80% stenosis carotid blood flow and energy dissipation are predicted to be reduced sufficiently to explain decreasing bruit intensity and possibly the clinical difficulty with obtaining accurate Doppler velocity measurements.

### Cerebral Blood Flow Reserve, Cerebrovascular Reactivity and Model Predictions

Both percent stenosis and mean systemic arterial pressure must be known for measured cerebral flow reserve and/or cerebrovascular reactivity results to be used with the model to estimate patient specific collateral vascular resistance values. The Rw curves in Figures 4 and 5 are blood flow reserve when cerebral flow is normal and when it is reduced at moderate or high-grade carotid stenosis. In asymptomatic patients with moderate carotid stenosis it is not practical or advisable to obtain Rw with carotid stump pressure measurements. If percent diameter carotid stenosis and cerebral reactivity or flow reserve are measured non-invasively the patient specific collateral vascular resistance curve, Rw, can be identified, as given in Figures 4 and 5, and reserve blood flow predicted. This model offers an alternative method of using cerebrovascular reactivity or auto regulatory vasodilation measurements to predict cameral blood flow reserve. Acetazolamide stimulation and either trans-cranial Doppler or Xenon CT or MRI cerebral blood flow measurements are the most commonly used methods. Grossman et al (22) reported an increase in cerebral blood flow of 40 ml/min/100g in normal subjects, almost double the normal value. When mean arterial pressure is less than 100 mmHg the model predicts a maximum reserve flow less than 50 ml/min/100g, as in Figure 5a. Patients with moderate to high-grade carotid stenosis or occlusion are reported to have ipsilateral flow reserve of 24% (23). Russell et al (24) report that prior to carotid endarterectomy patients had “reduced reserve” restored to “normal” after surgery. With patient specific cerebral blood flow reserve or reactivity data that includes mean arterial pressure and percent stenosis the model has the potential to predict patient specific flow curves.

### Critical Carotid Stenosis

The model solutions given in results predict critical carotid artery stenosis values 65.2% and 70.7% diameter stenosis. At higher degrees of stenosis carotid blood flow is not sufficient to maintain normal regional cerebral blood flow. These were calculated from the specific model equation solution when Rb = 1.0, Rw = ∞ and Q is normal 50ml/min/100g, or Rc = (Pa/50 - 1). When Pa = 87mmHg, Rc = 0.74 or 65.2% and when Pa = 100mmHg, Rc = 1.0 or 70.7%. These predictions are close to the measured values of 60% to 65% diameter stenosis in the Figure 8 study (19). These findings strongly support the functional relationship between Rc and % carotid stenosis given in Figure 3. In a clinical study of 665 patients with significant carotid stenosis (17) the mean arterial pressure Pa was 84mmHg. When Rw = ∞ this gives critical carotid stenosis of 63.6% (Rc = 0.68). Both of the clinically measured mean of the mean systemic arterial pressures, 84 mmHg and 87mmHg, seem low. A hypertensive mean systemic arterial pressure of 110 mmHg predicts critical stenosis to be 73.9%. This small variance in critical carotid artery percent diameter stenosis due to mean systemic arterial pressure not withstanding, the model illustrates the inadequacy of carotid blood flow alone to prevent cerebral ischemia at moderate levels of stenosis. Sufficient collateral flow is predicted to prevent cerebral ischemia when patient’s collateral resistance less than 1.0, Rw<1.0.

The quest to determine critical artery stenosis has a long and checkered history. Berger and Hwang’s (18) classic study of critical arterial stenosis was also based on an energy conservation-dissipation hemodynamic model. They assumed arterial stenosis pressure gradient to be proportional to the percent diameter stenosis and added a turbulent component. Animal peripheral artery model results matched the theory well, predicting an exponential vascular resistance curves. This finding is very similar to the second-order power function used herein for carotid stenosis vascular resistance. They recognized that measurement of the pressure gradient across a stenosis is not sufficient to define critical stenosis because of the dependency on flow rate. While this is correct and is predicted by the definition of vascular resistance, these results are not applicable to carotid artery stenosis because of parallel collateral flow coupled with a maintained relatively normal level of cerebral blood flow. Carotid artery stenosis does have critical values if collateral blood flow is not sufficient to maintain normal cerebral blood flow.

### The Model

The three-component steady flow hemodynamic model is based on the principal of conservation of energy. The total input energy per unit blood flow volume is the mean arterial pressure, Pa. Accurate measurements of arterial blood pressure are made with a calibrated manometer via catheter or needle. Mean systemic arterial and mean carotid artery stump pressures used in this study were made this way with the patients supine and zero pressure set at the carotid level which is small and neglected. This also sets a zero level for cerebral venous and cerebral fluid pressure. This means that the input energy is assumed completely dissipated by the three vascular resistance components. Carotid energy dissipation is primarily due to turbulent blood flow at the stenosis resulting in tissue deformation/vibration and heat in the neck. Collateral energy dissipation occurs at multiple locations in the circle of Willis, the contralateral carotid and the vertebral - basilar systems. The brain normally dissipates energy uniformly throughout the arteriolar microcirculation. In the absence of carotid stenosis, Rc = 0, all energy is dissipated in the brain. With carotid occlusion, Rc = ∞, all energy is dissipated by the brain and collateral components. For carotid stenosis between zero% and 100% the model solutions for cerebral and reserve blood flows are determined by the relative energy losses in each component.

The aim of this study was to determine if a basic hemodynamic model can predict the contributions of carotid and collateral blood flow to regional cerebral blood flow in patients with extra-cranial carotid stenosis. Hemodynamic models are used to simplify complex circulatory systems to the extent that approximate solutions can be obtained. The one-dimensional three-component model in Figure 1 is often used to represent a fluid mechanics or electrical circuit problem because the algebraic equations for the underlying physics laws are identical. When used to represent the cerebrovascular system this becomes complex because the carotid and collateral resistances, Rc and Rw, are patient specific non-linear variables and brain resistance Rb, is a linear variable. Anatomically the model is applicable to humans because the carotid, collateral and brain vascular systems are physically separate while arterially connected. Solutions were possible in this study because carotid stenosis vascular resistance was assumed to be proportional to percent area carotid stenosis, as given in Figure 3, and model solutions predict patient specific values if collateral vascular resistance. The solutions for cerebral blood flow, cerebral blood flow reserve, energy dissipation, critical carotid stenosis and cervical bruit are predictions of the how and why clinical events and observations. Perhaps future complex computational solutions using modern blood flow equations will provide a more accurate and/or precise estimate of reality but this simple modified Bernoulli energy model appears to do fairly well.

## Data Availability

All data is published and referenced

## Acknowledgements

Sarah Ellen my wife of 60 years was once again a trooper by typing and editing the drafts and supporting this effort to generate one more paper. Thanks to Sujal Dave at NCSU who did an excellent job formatting the figures. In memory of my fishing and hunting buddies Jim Connell and Tommy Thompson.

